# Economic burden of cancer and cardiovascular disease mortality among working-age Europeans: A lifecycle modelling study

**DOI:** 10.64898/2026.02.13.26346233

**Authors:** Paul A. Hanly, Marta Ortega–Ortega, Yek-Ching Kong, Marianna De Camargo Cancela, Isabelle Soerjomataram

## Abstract

**Objectives:** Non-communicable diseases (NCDs) account for almost 90% of deaths in Europe, yet comparative estimates of the productivity costs associated with premature NCD mortality across diseases and countries remain limited. This study estimates and compares productivity losses attributable to cardiovascular disease (CVD) and cancer mortality among working-age populations across Europe. Population-based data were used to estimate productivity costs for CVD and cancer deaths across 30 European countries. Sex- and age-specific mortality data for 2021 were obtained from the World Health Organization Mortality Database. Economic data, including wages, unemployment rates, and labour force participation rates, were sourced from Eurostat. Productivity losses were valued using a human capital approach incorporating an age-transition lifecycle simulation model that adjusts for lifetime wage trajectories and labour market dynamics. Costs were discounted at 3.5%. Total productivity losses from cancer and CVD mortality in working-age populations were estimated at €195.7 billion, equivalent to 1.24% of European GDP. Cancer accounted for 62.5% (€122.2 billion) of total productivity losses, while CVD accounted for 37.5% (€73.5 billion). Total CVD-related productivity costs exceeded cancer-related costs in Central and Eastern Europe, whereas cancer productivity costs were higher in Western, Northern, and Southern Europe. Mean productivity costs per death were higher for CVD (€219,848; 95% CI 165,241–270,247) than for cancer (€217,744; 95% CI 166,554–273,144). A larger gender gap was observed for CVD mortality, with a male-to-female cost ratio of 2.5 compared with 1.6 for cancer. Productivity losses associated with premature cancer and CVD mortality represent a substantial economic burden across Europe, with pronounced variation by disease, region, and sex. These findings provide comparative, cross-country estimates of the human capital costs associated with major NCD causes of death.

## Introduction

The European economy is currently the lowest growing region globally due to ageing populations, stagnating productivity and, reducing competitiveness [1]. The stock of human capital is essential to long-term economic growth, and disease-related premature mortality reduces human capital and productivity, lowers gross domestic product (GDP) and leads to diminished growth prospects over time [2–3]. This impact is particularly acute in the case of populations with ageing workforces who are burdened by premature mortality associated with Non-Communicable Diseases (NCDs) [4].

NCDs are a leading cause of death globally, accounting for 74% of all deaths [5]. While NCDs are generally associated with older age groups, 17 million NCDs-related deaths were reported among those under 70 years of age [6]. In Europe, the mortality burden is even greater, where 90% of deaths are attributable to NCDs [5]. The leading causes of death from NCDs are cardiovascular diseases (CVDs) (43%) and cancer (23%), together accounting for two-thirds of deaths across Europe [6]. This burden is forecasted to grow over time commensurate with the aging of populations across Europe and the growing prevalence of unhealthy lifestyles [7]. These chronic illnesses can greatly impact productivity due to reduced labour force participation, presenteeism, absence from work, long-term disability and, the focus of this study – deaths in the working age population [2,3,8].

Productivity costs associated with death account for the largest proportion of indirect costs associated with CVD [9] and cancer [10], with estimates of 17% and 25% of the total burden for CVD and cancer respectively. While studies focused on the health burden of diseases, including CVD and cancer, are widespread, disease-related productivity cost studies are less prevalent, and tend to focus on a single NCD if multinational in nature [11, 12]. Rarely do studies attempt to evaluate productivity costs across more than one disease, and across multiple countries.

Precise and reliable productivity cost estimates from deaths due to NCDs are vital to inform policy decisions aimed at reducing their burden, particularly as these costs can alternatively be viewed as potential cost savings arising from a reduction in deaths. The Sustainable Development Goal (SDG) target 3.4 is to reduce premature mortality from non-communicable diseases (NCDs) by a third by 2030 relative to 2015 levels through prevention and treatment [13]. Standardized and comparable multinational cost estimates of NCDs are consequently vital in establishing a baseline for evaluating the potential economic benefits of this goal, and to aid in guiding the allocation of government resources and informing priorities for cancer and CVD control in Europe. Accordingly, the aim of this study is to apply standardised economic approaches to population-based data to estimate productivity costs associated with CVD and cancer deaths in the working age population across Europe as a whole, by sex, age, country and region.

## Materials and Methods

### Data Sources

The analysis was based on a combination of health and economic data extracted for 30 European countries (the 27 European Union countries, plus Norway, Switzerland and Iceland, listed in S1 Table) and classified into four European regions (Northern, Southern, Central-Eastern (C-E) and, Western Europe).

The sex- and age-specific (10-year age bands from 15-24 to 65-74) number of deaths in 2021 (or closest year available) by country for both malignant neoplasms, ICD-10 codes: C00-C97 (hereafter referred to as cancer), and cardiovascular diseases, ICD-10 codes: I00-I99 (CVD), were obtained from the World Health Organization Mortality Database [14]. The decision to extend the cut-off age of the working population to 74 years (beyond the traditional cutoff at 65 years) was based on evidence of a growing proportion of the workforce engaging in the labour force beyond the traditional retirement age (between 3% and 22% in Europe in 2021 [15]).

Economic data included wages, unemployment rates and labour force participation rates. Country, age and sex-specific annual wage data were obtained from the Eurostat Structure of Earnings Survey for 2018 [15], defined as the annual gross earnings (for both part- and full-time jobs) for industry, construction and services in 2018. Data were inflated to 2021 prices based on the calculated wage growth per country between the 2010 Structure of Earnings Survey and the 2018 survey [15]. Country, age and sex-specific data on unemployment and labour force participation were extracted from Eurostat and based on the European Labour Force Survey 2021 [15]. Where economic data was missing for a specific country, we used the geographic region average.

### Estimation methods

We estimated productivity costs for the working population using the well-established Human Capital Approach. According to this theory, premature mortality arises when an individual dies from a disease prior to retirement and is therefore removed from the work force, impacting the potential future production in an economy. The productivity lost to the economy arising from this potential stream of labour output between age of death and retirement age can be valued using the individual’s working wage (an indirect measure of their productivity) [16].

We developed a simulation model to estimate the productivity cost from cancer and CVD deaths. We first calculated years of potential productive life lost (YPPLL) by sex and age-group using the WHO Standard Life Tables, defined as the number of years of life lost between the midpoint of each age group at death and 74 years of age (YPPLL = life expectancy by age group up to 74 - age at death). Valuation of productivity costs was based on multiplying YPPLL by country, sex and age-specific annual wages until age 74, and adjusting for unemployment, labour force participation and wage growth rates. In a modification to traditional approaches, our productivity costs for each death were aggregated to include future wage progression. For example, an individual who dies of cancer aged 15-29 would potentially have progressed through the labour market had they lived, entering successively higher wage groups with age and experience. This hypothetical progression is incorporated through lifecycle wage progression in the model. Future wage growth was estimated based on average country-specific wage growth from 2010 to 2018, with data obtained from the Eurostat Structure of Earnings Survey [15] for both years. Future costs were discounted at 3.5% per annum.

Sensitivity analysis included a gender-neutral estimate where the average wage, instead of sex-specific wages, was applied to both sexes, a cut-off age of 65, Purchasing Power Parity (PPP)-adjusted wages to account for cost of living differences across the European regions, in addition to the use of lower (0%) and upper (6%) bound estimates for the discount rate.

## Results

In 2021, cancer accounted for more deaths (0·58 million deaths versus 0·42 million deaths) (S2 Table), more YPPLL (7·04 million YPPLL versus 4·78 million YPPLL) (S3 Table) and a higher YPPLL per death ratio (12·2 YPPLL per death versus 11·4 YPPLL per death) (S3 Table) than CVD amongst working age people in Europe. Western Europe accounted for the largest cancer burden by region at 40% of deaths and 40% of YPPLL. Only C-E Europe, out of the four European regions, exhibited a higher CVD burden across these health metrics.

### Productivity costs associated with cancer and CVD deaths (Total, per death and per GDP)

The total cost of lost productivity associated with cancer and CVD deaths for Europe in 2021 was €195·7 billion, of which cancer accounted for the larger proportion (62·5%; CVD 37·5%) (Table 1 and Figs 1, 2 and 3). On a regional basis, the cost burden exhibited a distinct East-West regional divide. CVD costs tended to be larger in C-E Europe (Cancer/CVD ratio: 0·93) while cancer productivity costs exceeded those of CVD across Western (ratio: 1·74) and Southern Europe (ratio: 2·04). In Northern Europe, the cost burden was mixed, where four countries (Estonia, Finland, Latvia and Lithuania) experienced a higher CVD cost burden than cancer, although total cancer costs remained higher overall (ratio: 1·58).

**Table 1:** Total productivity cost (‘000,000) for cancer and cardiovascular disease (CVD) deaths among those aged 15-74 in Europe in 2021, overall and by sex.

| Country/R<br>egion | Cancer premature mortality cost (€) |  |  |  |  |  |  | CVD premature mortality cost (€) |  |  |  |  |  |  | Combined premature<br>mortality cost (€) |  |
| --- | --- | --- | --- | --- | --- | --- | --- | --- | --- | --- | --- | --- | --- | --- | --- | --- |
|  | Male cost<br>(€) | % | Female<br>cost (€) | % | Male/Femal<br>e ratio | Total cost<br>(€) | % | Male cost<br>(€) | % | Female<br>cost (€) | % | Male/Femal<br>e ratio | Total cost<br>(€) | % | Total<br>Cancer<br>and CVD<br>Cost (€) | % |
| Bulgaria | 318.4 |  | 233.1 |  | 1.37 | 551.5 |  | 834.8 |  | 306.9 |  | 2.72 | 1,141.7 |  | 1,693.1 |  |
| Czechia | 868.0 |  | 509.8 |  | 1.70 | 1,377.8 |  | 981.8 |  | 234.7 |  | 4.18 | 1,216.5 |  | 2,594.3 |  |
| Hungary | 841.4 |  | 566.7 |  | 1.48 | 1,408.1 |  | 1,157.6 |  | 329.9 |  | 3.51 | 1,487.5 |  | 2,895.7 |  |
| Poland | 2,440.3 |  | 1,542.0 |  | 1.58 | 3,982.3 |  | 3,096.6 |  | 706.6 |  | 4.38 | 3,803.2 |  | 7,785.5 |  |
| Romania | 1,333.0 |  | 738.5 |  | 1.81 | 2,071.4 |  | 1,872.9 |  | 501.2 |  | 3.74 | 2,374.1 |  | 4,445.5 |  |
| Slovakia | 431.9 |  | 256.2 |  | 1.69 | 688.0 |  | 642.2 |  | 152.3 |  | 4.22 | 794.5 |  | 1,482.5 |  |
| **C-E**<br>**Europe** | **6,233.0** | **8.2** | **3,846.2** | **8.3** | **1.62** | **10,079.3** | **8.2** | **8,585.9** | **16.3** | **2,231.5** | **10.7** | **3.85** | **10,817.4** | **14.7** | **20,896.7** | **10.7** |
| Denmark | 1,491.7 |  | 1,006.4 |  | 1.48 | 2,498.1 |  | 858.9 |  | 250.9 |  | 3.42 | 1,109.8 |  | 3,607.8 |  |
| Estonia | 106.5 |  | 71.4 |  | 1.49 | 177.9 |  | 146.9 |  | 38.3 |  | 3.83 | 185.2 |  | 363.1 |  |
| Finland | 900.7 |  | 181.1 |  | 4.97 | 1,081.8 |  | 941.7 |  | 224.3 |  | 4.20 | 1,166.0 |  | 2,247.9 |  |
| Iceland | 91.1 |  | 68.5 |  | 1.33 | 159.6 |  | 65.9 |  | 16.7 |  | 3.96 | 82.6 |  | 242.2 |  |
| Ireland | 1,019.4 |  | 642.5 |  | 1.59 | 1,661.9 |  | 776.9 |  | 160.7 |  | 4.83 | 937.6 |  | 2,599.6 |  |
| Latvia | 154.9 |  | 109.4 |  | 1.42 | 264.2 |  | 367.2 |  | 103.3 |  | 3.56 | 470.5 |  | 734.8 |  |
| Lithuania | 200.7 |  | 148.3 |  | 1.35 | 349.0 |  | 353.0 |  | 99.4 |  | 3.55 | 452.4 |  | 801.4 |  |
| Norway | 995.4 |  | 729.4 |  | 1.36 | 1,724.8 |  | 566.4 |  | 151.3 |  | 3.74 | 717.7 |  | 2,442.5 |  |
| Sweden | 1,319.4 |  | 1,079.7 |  | 1.22 | 2,399.1 |  | 1,114.8 |  | 304.5 |  | 3.66 | 1,419.3 |  | 3,818.4 |  |
| **Northern**<br>**Europe** | **6,279.8** | **8.3** | **4,036.6** | **8.7** | **1.56** | **10,316.4** | **8.4** | **5,191.8** | **9.9** | **1,349.4** | **6.5** | **3.85** | **6,541.2** | **8.9** | **16,857.6** | **8.6** |
| Croatia | 309.1 |  | 193.4 |  | 1.60 | 502.5 |  | 274.8 |  | 69.0 |  | 3.98 | 343.8 |  | 846.3 |  |
| Cyprus | 84.0 |  | 51.4 |  | 1.63 | 135.4 |  | 88.0 |  | 21.1 |  | 4.18 | 109.1 |  | 244.4 |  |
| Greece | 1,236.3 |  | 531.9 |  | 2.32 | 1,768.3 |  | 1,155.7 |  | 190.5 |  | 6.07 | 1,346.2 |  | 3,114.5 |  |
| Italy | 9,469.2 |  | 5,000.8 |  | 1.89 | 14,470.1 |  | 5,533.1 |  | 1,102.9 |  | 5.02 | 6,636.0 |  | 21,106.1 |  |
| Malta | 50.2 |  | 27.0 |  | 1.86 | 77.1 |  | 52.1 |  | 11.0 |  | 4.72 | 63.1 |  | 140.3 |  |
| Portugal | 1,284.4 |  | 661.4 |  | 1.94 | 1,945.8 |  | 648.7 |  | 173.8 |  | 3.73 | 822.6 |  | 2,768.4 |  |
| Slovenia | 257.0 |  | 163.8 |  | 1.57 | 420.8 |  | 145.0 |  | 33.1 |  | 4.38 | 178.1 |  | 598.9 |  |
| Spain | 6,052.0 |  | 3,370.9 |  | 1.80 | 9,422.9 |  | 3,718.8 |  | 844.1 |  | 4.41 | 4,562.9 |  | 13,985.8 |  |
| **Southern Europe** | **18,742.1** | **24.7** | **10,000.8** | **21.6** | **1.87** | **28,742.9** | **23.5** | **11,616.2** | **22.1** | **2,445.6** | **11.7** | **4.75** | **14,061.8** | **19.1** | **42,804.7** | **21.9** |
| Austria | 1,865.8 |  | 1,087.4 |  | 1.72 | 2,953.2 |  | 1,353.9 |  | 315.2 |  | 4.30 | 1,669.1 |  | 4,622.3 |  |
| Belgium | 1,953.6 |  | 1,410.1 |  | 1.39 | 3,363.7 |  | 1,043.2 |  | 358.7 |  | 2.91 | 1,401.9 |  | 4,765.6 |  |
| France | 11,602.6 |  | 7,098.5 |  | 1.63 | 18,701.0 |  | 4,989.8 |  | 1,377.7 |  | 3.62 | 6,367.5 |  | 25,068.5 |  |
| Germany | 22,089.6 |  | 13,716.4 |  | 1.61 | 35,805.9 |  | 16,120.0 |  | 11,751.6 |  | 1.37 | 27,871.6 |  | 63,677.5 |  |
| Luxembourg | 129.6 |  | 87.3 |  | 1.48 | 217.0 |  | 77.4 |  | 19.4 |  | 3.99 | 96.8 |  | 313.8 |  |
| Switzerland | 2,821.1 |  | 1,735.2 |  | 1.63 | 4,556.2 |  | 1,573.6 |  | 333.6 |  | 4.72 | 1,907.2 |  | 6,463.5 |  |
| Netherlands | 4,287.8 |  | 3,203.1 |  | 1.34 | 7,490.9 |  | 2,035.2 |  | 690.2 |  | 2.95 | 2,725.4 |  | 10,216.3 |  |
| **Western Europe** | **44,750.1** | **58.9** | **28,337.9** | **61.3** | **1.58** | **73,088.0** | **59.8** | **27,193.1** | **51.7** | **14,846.5** | **71.1** | **1.83** | **42,039.5** | **57.2** | **115,127.5** | **58.8** |
| **Europe Total** | **76,005.0** | **100.0** | **46,221.5** | **100.0** | **1.64** | **122,226.6** | **100.0** | **52,586.9** | **100.0** | **20,873.0** | **100.0** | **2.52** | **73,459.9** | **100.0** | **195,686.5** | **100.0** |

**Fig 1:**
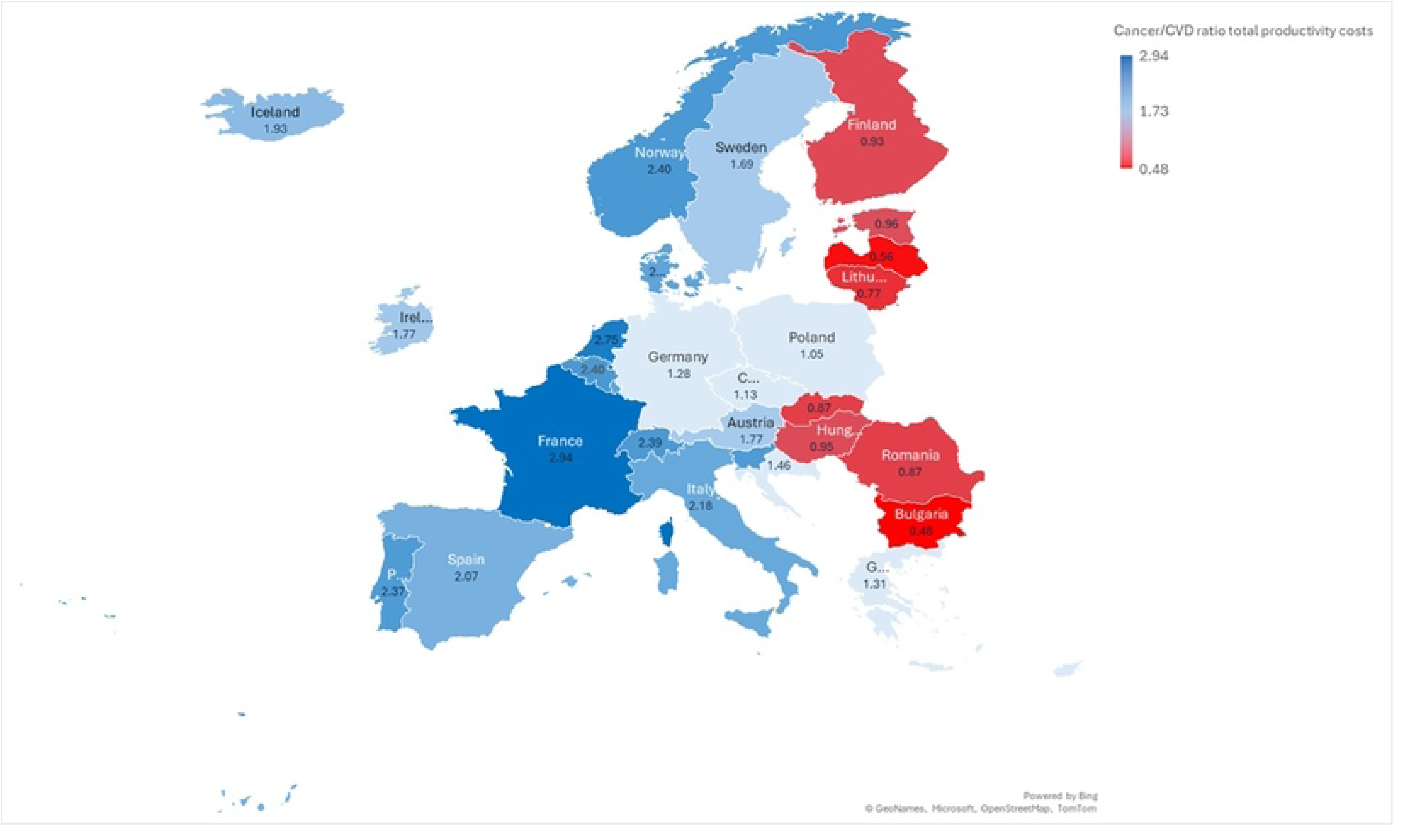
Cancer/cardiovascular disease (CVD) total productivity cost ratio among those aged 15-74 in Europe in 2021

**Fig 2:**
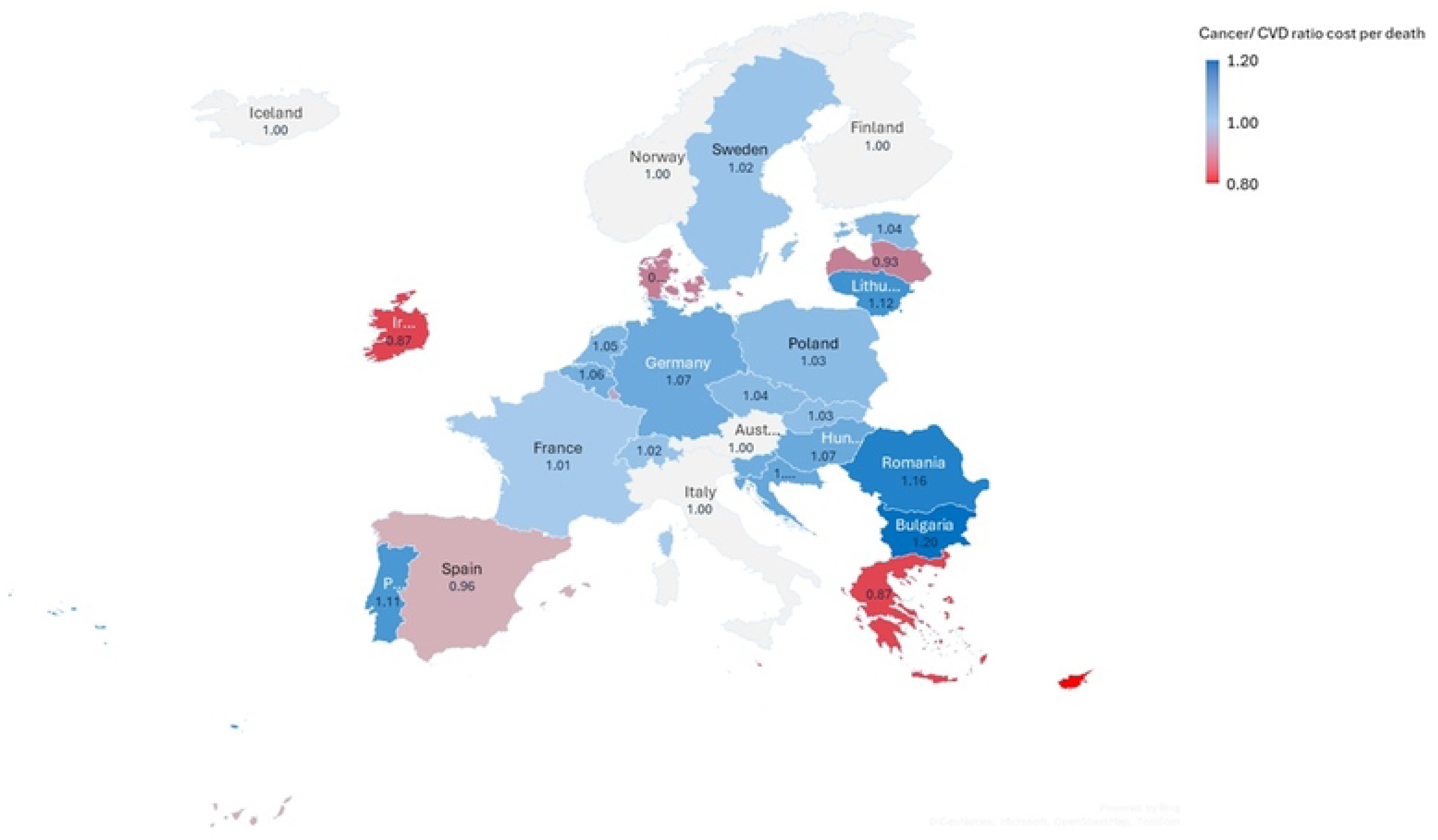
Cancer/cardiovascular disease (CVD) productivity cost per death ratio among those aged 15-74 in Europe in 2021

**Fig 3:**
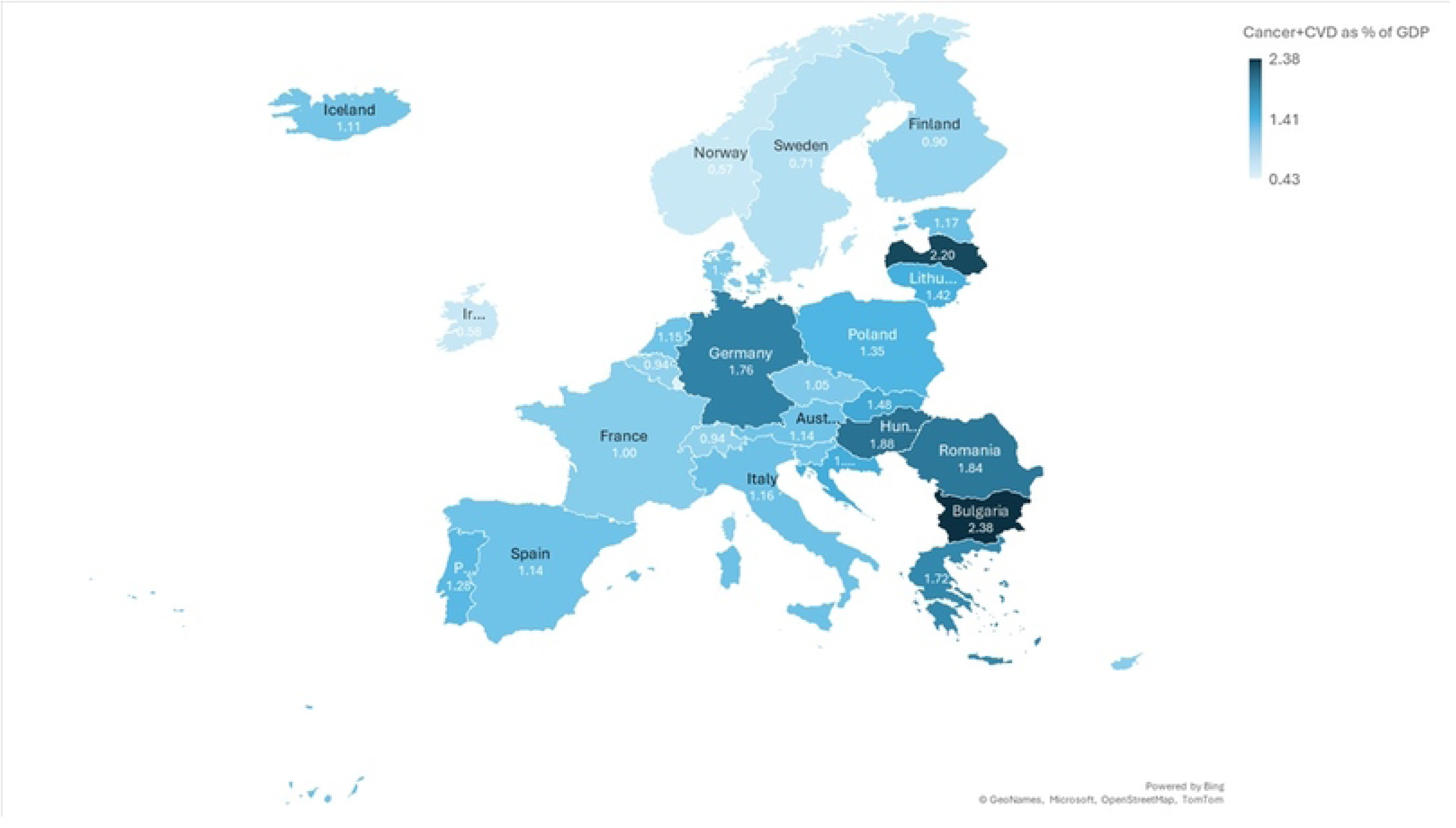
Total productivity cost for cancer and cardiovascular disease (CVD) combined as a % of Gross Domestic Product among those aged 15-74 in Europe in 2021

For both diseases, Western and Southern Europe experienced the highest cost burden with €73·1 billion and €28·7 billion from cancer deaths (59·8% and 23·5% of the European total), and €42·0 billion and €14·0 billion from CVD deaths (57·2% and 19·1% of the European total). The third and fourth ranked regions for cancer are Northern (8·4%) and C-E Europe (8·2%), however for CVD, C-E Europe ranked third (14·7%) and Northern Europe fourth (8·9%). The most populous countries experienced the highest cost burden, although the ranking changed somewhat between cancer (Germany: €35·8 billion, France: €18·7 billion, Italy: €14·5 billion) and CVD (Germany: €27·9 billion, Italy: €6·6 billion, France: €6·4 billion).

In Table 2, total CVD costs per death (€219,848; 95% Confidence Interval [CI] 165,241-270,247) were greater than cancer costs per death (€217,744, 95% CI 166,554-273,144) across Europe as a whole, and in two of the four regions (C-E Europe and Western Europe). In addition, the highest relative burden for CVD compared to cancer was exhibited across Europe’s periphery (Ireland, Denmark, Latvia, Greece, Cyprus, Malta and Spain). Rather than the cost burden being dominated by the most populous countries (Germany, France and Italy), cost per death revealed Switzerland (cancer: €611,567; CVD: €600,703), Iceland: (cancer: €555,986; CVD: €554,278) and Luxembourg (cancer: €431,317; CVD: €446,217) as the highest cost countries for cancer and CVD, and Bulgaria as the lowest (cancer: €50,121; CVD: €41,692).

**Table 2:**
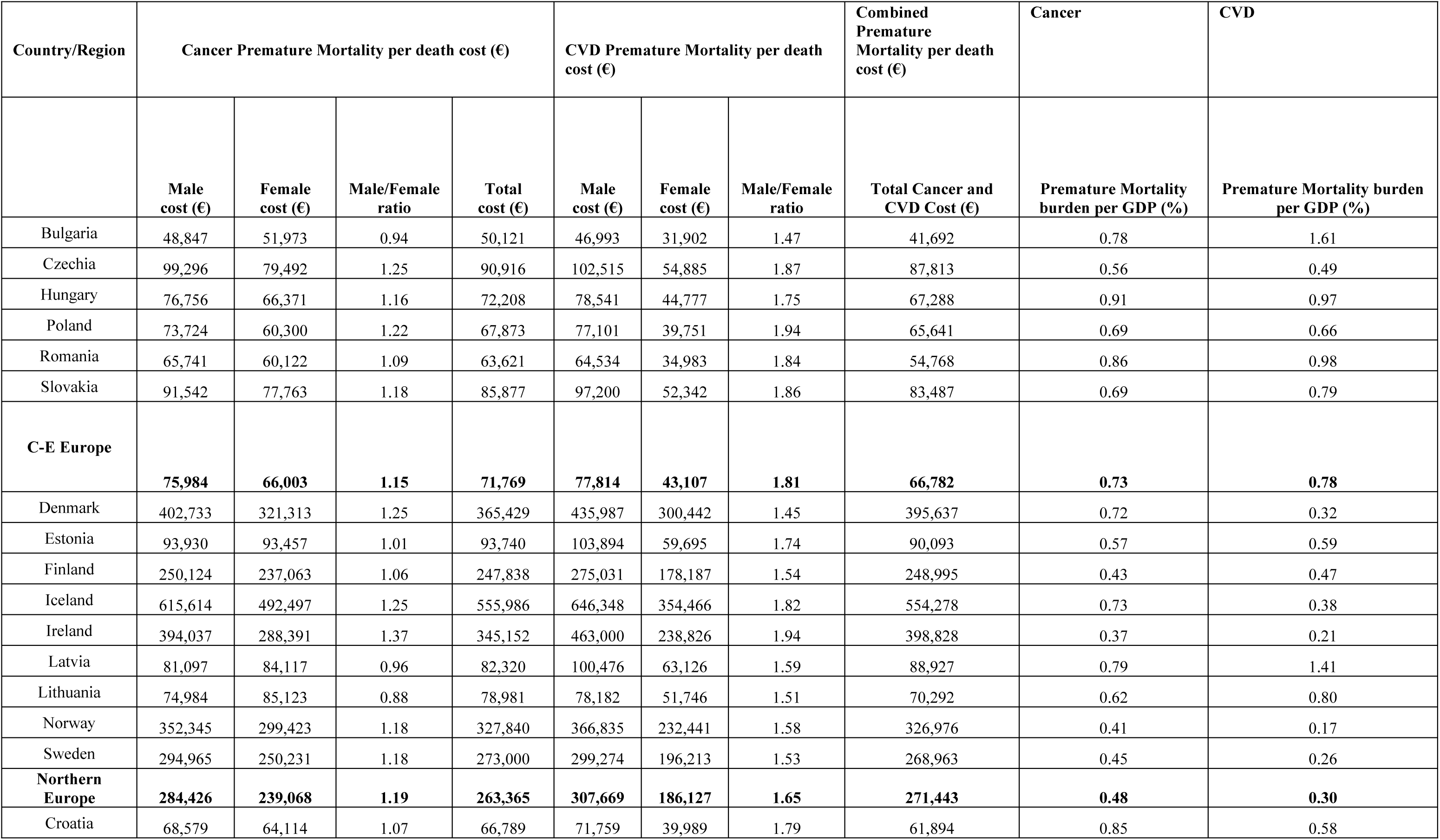
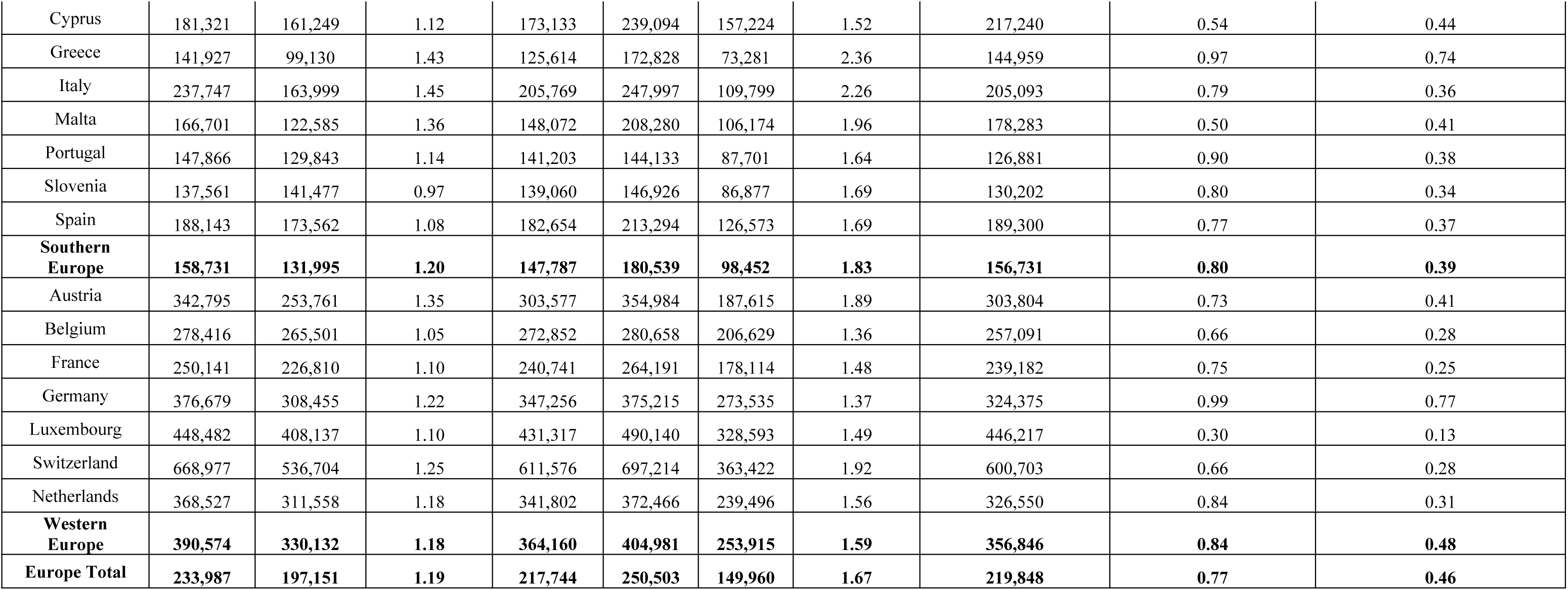
Average productivity cost per death for cancer and cardiovascular disease (CVD) and total productivity costs as a percentage of GDP among those aged 15-74 in Europe in 2021, overall and by sex.

| Country/Region | Cancer Premature Mortality per death cost (€) |  |  |  | CVD Premature Mortality per death cost (€) |  |  | Combined Premature Mortality per death cost (€) | Cancer | CVD |
| --- | --- | --- | --- | --- | --- | --- | --- | --- | --- | --- |
|  | Male cost (€) | Female cost (€) | Male/Female ratio | Total cost (€) | Male cost (€) | Female cost (€) | Male/Female ratio | Total Cancer and CVD Cost (€) | Premature Mortality burden per GDP (%) | Premature Mortality burden per GDP (%) |
| Bulgaria | 48,847 | 51,973 | 0.94 | 50,121 | 46,993 | 31,902 | 1.47 | 41,692 | 0.78 | 1.61 |
| Czechia | 99,296 | 79,492 | 1.25 | 90,916 | 102,515 | 54,885 | 1.87 | 87,813 | 0.56 | 0.49 |
| Hungary | 76,756 | 66,371 | 1.16 | 72,208 | 78,541 | 44,777 | 1.75 | 67,288 | 0.91 | 0.97 |
| Poland | 73,724 | 60,300 | 1.22 | 67,873 | 77,101 | 39,751 | 1.94 | 65,641 | 0.69 | 0.66 |
| Romania | 65,741 | 60,122 | 1.09 | 63,621 | 64,534 | 34,983 | 1.84 | 54,768 | 0.86 | 0.98 |
| Slovakia | 91,542 | 77,763 | 1.18 | 85,877 | 97,200 | 52,342 | 1.86 | 83,487 | 0.69 | 0.79 |
| **C-E Europe** | **75,984** | **66,003** | **1.15** | **71,769** | **77,814** | **43,107** | **1.81** | **66,782** | **0.73** | **0.78** |
| Denmark | 402,733 | 321,313 | 1.25 | 365,429 | 435,987 | 300,442 | 1.45 | 395,637 | 0.72 | 0.32 |
| Estonia | 93,930 | 93,457 | 1.01 | 93,740 | 103,894 | 59,695 | 1.74 | 90,093 | 0.57 | 0.59 |
| Finland | 250,124 | 237,063 | 1.06 | 247,838 | 275,031 | 178,187 | 1.54 | 248,995 | 0.43 | 0.47 |
| Iceland | 615,614 | 492,497 | 1.25 | 555,986 | 646,348 | 354,466 | 1.82 | 554,278 | 0.73 | 0.38 |
| Ireland | 394,037 | 288,391 | 1.37 | 345,152 | 463,000 | 238,826 | 1.94 | 398,828 | 0.37 | 0.21 |
| Latvia | 81,097 | 84,117 | 0.96 | 82,320 | 100,476 | 63,126 | 1.59 | 88,927 | 0.79 | 1.41 |
| Lithuania | 74,984 | 85,123 | 0.88 | 78,981 | 78,182 | 51,746 | 1.51 | 70,292 | 0.62 | 0.80 |
| Norway | 352,345 | 299,423 | 1.18 | 327,840 | 366,835 | 232,441 | 1.58 | 326,976 | 0.41 | 0.17 |
| Sweden | 294,965 | 250,231 | 1.18 | 273,000 | 299,274 | 196,213 | 1.53 | 268,963 | 0.45 | 0.26 |
| **Northern Europe** | **284,426** | **239,068** | **1.19** | **263,365** | **307,669** | **186,127** | **1.65** | **271,443** | **0.48** | **0.30** |
| Croatia | 68,579 | 64,114 | 1.07 | 66,789 | 71,759 | 39,989 | 1.79 | 61,894 | 0.85 | 0.58 |
| Cyprus | 181,321 | 161,249 | 1.12 | 173,133 | 239,094 | 157,224 | 1.52 | 217,240 | 0.54 | 0.44 |
| Greece | 141,927 | 99,130 | 1.43 | 125,614 | 172,828 | 73,281 | 2.36 | 144,959 | 0.97 | 0.74 |
| Italy | 237,747 | 163,999 | 1.45 | 205,769 | 247,997 | 109,799 | 2.26 | 205,093 | 0.79 | 0.36 |
| Malta | 166,701 | 122,585 | 1.36 | 148,072 | 208,280 | 106,174 | 1.96 | 178,283 | 0.50 | 0.41 |
| Portugal | 147,866 | 129,843 | 1.14 | 141,203 | 144,133 | 87,701 | 1.64 | 126,881 | 0.90 | 0.38 |
| Slovenia | 137,561 | 141,477 | 0.97 | 139,060 | 146,926 | 86,877 | 1.69 | 130,202 | 0.80 | 0.34 |
| Spain | 188,143 | 173,562 | 1.08 | 182,654 | 213,294 | 126,573 | 1.69 | 189,300 | 0.77 | 0.37 |
| **Southern**<br>**Europe** | **158,731** | **131,995** | **1.20** | **147,787** | **180,539** | **98,452** | **1.83** | **156,731** | **0.80** | **0.39** |
| Austria | 342,795 | 253,761 | 1.35 | 303,577 | 354,984 | 187,615 | 1.89 | 303,804 | 0.73 | 0.41 |
| Belgium | 278,416 | 265,501 | 1.05 | 272,852 | 280,658 | 206,629 | 1.36 | 257,091 | 0.66 | 0.28 |
| France | 250,141 | 226,810 | 1.10 | 240,741 | 264,191 | 178,114 | 1.48 | 239,182 | 0.75 | 0.25 |
| Germany | 376,679 | 308,455 | 1.22 | 347,256 | 375,215 | 273,535 | 1.37 | 324,375 | 0.99 | 0.77 |
| Luxembourg | 448,482 | 408,137 | 1.10 | 431,317 | 490,140 | 328,593 | 1.49 | 446,217 | 0.30 | 0.13 |
| Switzerland | 668,977 | 536,704 | 1.25 | 611,576 | 697,214 | 363,422 | 1.92 | 600,703 | 0.66 | 0.28 |
| Netherlands | 368,527 | 311,558 | 1.18 | 341,802 | 372,466 | 239,496 | 1.56 | 326,550 | 0.84 | 0.31 |
| **Western**<br>**Europe** | **390,574** | **330,132** | **1.18** | **364,160** | **404,981** | **253,915** | **1.59** | **356,846** | **0.84** | **0.48** |
| **Europe Total** | **233,987** | **197,151** | **1.19** | **217,744** | **250,503** | **149,960** | **1.67** | **219,848** | **0.77** | **0.46** |

Overall, the total cost of lost productivity in the working population due to CVD and cancer in Europe was equivalent to 1·24% of the combined GDP in 2021 (ranging from 0·4% in Luxembourg to 2·4% in Bulgaria) as shown in Fig 3. The highest cost relative to regional GDP was observed in C-E Europe (1·5%) which included three of the five countries with the highest cost burden relative to GDP in Europe (Bulgaria, Hungary and Romania). This was followed by Western Europe (combined: 1·3%), Southern Europe (combined: 1·2%) and Northern Europe (combined: 0·8%).

### Male versus female productivity cost burden of Cancer and CVD deaths

As shown in Table 1, total male productivity costs in European countries were higher than female costs for cancer (Male costs €76·0 billion vs. female costs €46·2 billion (M/F ratio: 1·6)) and for CVD (Male costs €52·6 billion vs. female costs €20·9 billion (M/F ratio: 2·5)). This difference was almost twice as much on average across all regions.

Productivity cost per death revealed far less divergence in male/female cost ratios (Table 2). For cancer, the overall male/female per death cost ratio was 1·2 (Male cost €233,987 vs. female costs €197,151), and for CVD it was 1·7 (Male cost €250,503 vs. female costs €149,960).

### Productivity cost burden of Cancer and CVD by age

Fig 4 shows the breakdown of total productivity costs by age for cancer and CVD. The 50-64 age group exhibited the highest cost for both cancer (€88.6 billion, 72.5%) and for CVD (€51.2 billion, 69.7%). The greatest divergence between cancer and CVD costs by age arose in the 15-29 age group, where cancer costs were 1.9 times higher compared to an average 1.7 times difference across all age groups.

**Figure 4:**
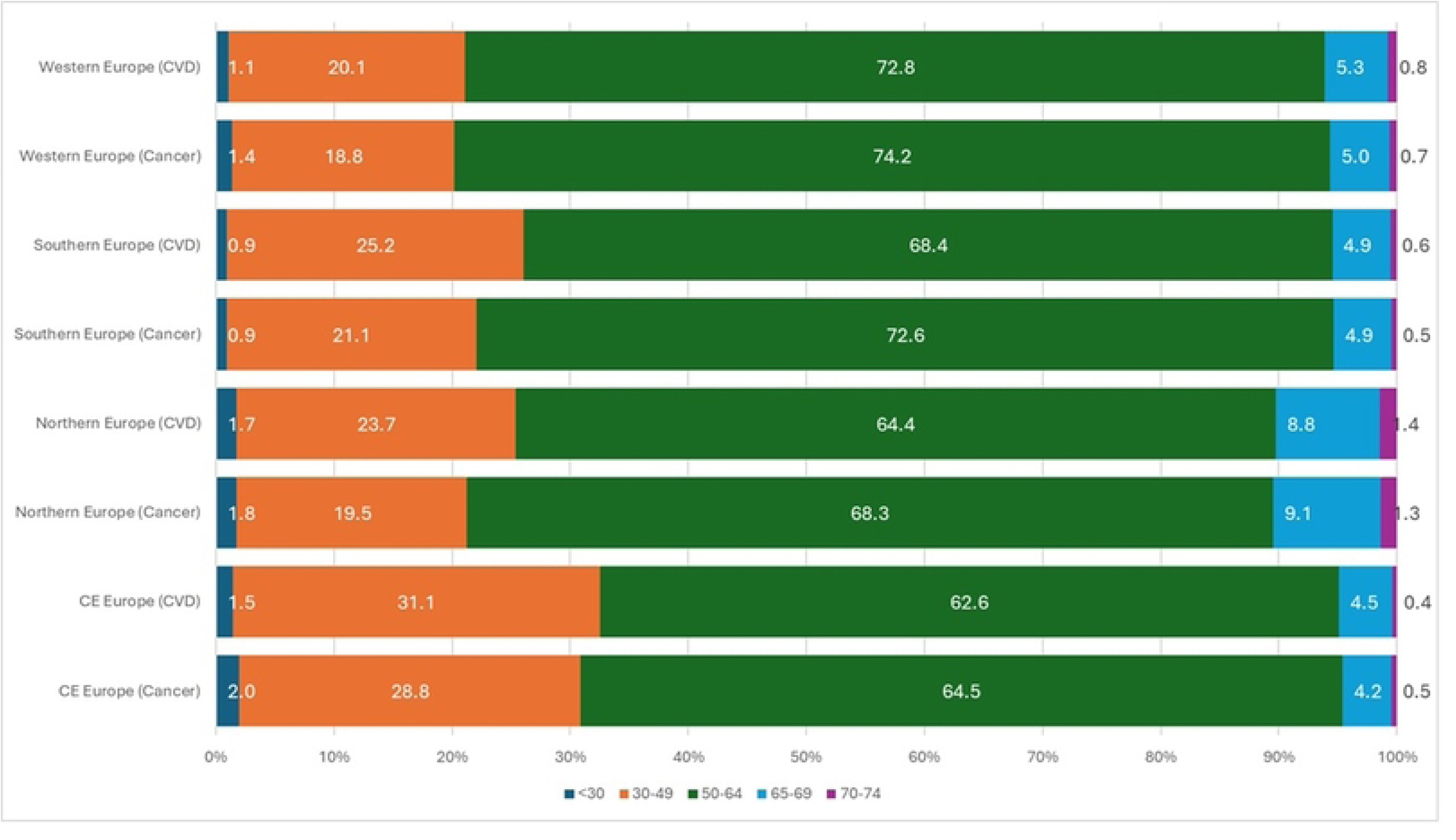
Total productivity cost (‘000,000) for cancer and cardiovascular disease (CVD) by age group among those aged 15-74 in Europe in 2021

### Sensitivity analysis

The use of gender-neutral wages reduced total productivity costs for cancer and CVD by a relatively small proportion (−0·88% and -1·24% respectively), however the male/female productivity cost burden ratio decreased by 16% across Europe for cancer and 14% for CVD (S4 Table). Introducing a cut-off age of 65 reduced productivity costs by almost a half for cancer and CVD (to €63.2 billion and €37.8 billion respectively). Purchasing Power Parity (PPP) adjusted estimates were 7·8% higher than the reference case for cancer and 13·0% for CVD, with the highest increase experienced in CE Europe (+83%). Discount rates of 0% and 6% resulted in lower bound estimates (€99·2 billion for cancer and €59·8 billion for CVD) and upper bound estimates (€167·0 billion for cancer and €99·9 billion for CVD) of total productivity costs.

## Discussion

### The economic burden of NCDs across Europe

Rarely are comparative assessments of the economic costs of two or more diseases undertaken across Europe. We estimated that lost productivity for cancer and CVD amongst those of working age in Europe amounted to €195·7 billion (1·24% of the European GDP).

Our estimates can alternatively be viewed as an indication of the potential savings from reducing deaths in the working age population from an economic perspective.

Our study reveals that the cost of lost productivity from cancer deaths tend to be larger across Europe as a whole (1·7 times greater), and across Southern (2·0 times), Western (1·7 times), and Northern Europe (1·6 times), despite traditional health burden metrics showing CVD as the leading cause of NCD death in Europe across all age groups [14]. A key reason for this is the comparatively higher lifetime risk of cancer at younger ages, while for CVD, the lifetime risk is higher at older ages [17]. This is evidenced by the higher productivity costs estimated among younger age groups from cancer in this study.

Evidence suggests that a significant proportion of NCDs are preventable through the reduction of behavioural risk factors [18]. These have been targeted by the WHO Global Action Plan for the Prevention and Control of Non-Communicable Diseases 2013-2030 [19] through a roadmap of multisectoral collaboration and the implementation of evidence-based interventions. The WHO European Action Plan for NCDs 2016-2025 [20] built on this with priorities focused on the European context, however, further investment is required, and our estimates suggest the potential economic value of this to European countries.

Progress has been made, for example, with regard to reduced tobacco use through the WHO Framework Convention on Tobacco Control (FCTC) [21] and reduced sugar consumption through fiscal policies including taxes on sugar-sweetened beverages and unhealthy foods [22]. However, more can be done, with greater resources necessary for tobacco control and greater scope for fiscal policy targeting across Europe.

Initiatives such as Europe’s Beating Cancer Plan [23] show evidence of progress in this area, however, key weaknesses remain. Europe has historically emphasised treatment rather than prevention for cancer care [20]. Evidence demonstrates that intervening at an early stage and prioritising disease prevention is more cost-effective compared to treating advanced-stage NCDs such as cancer [24]. Prioritising funds and advancing policies in disease prevention therefore exhibits significant scope to yield substantial economic benefits, particularly across Western, Northern and Southern Europe where the cancer burden exceeds that of CVD by a factor of 1·8 in the working age population, and where prevention financing has traditionally been low (5.1% of total health budget) [25]. While advocating for higher investments into cancer prevention efforts is important and potentially cost-saving, this should not be at the expense of continued resource allocations towards ensuring timely access to life-saving cancer treatments to patients and their families.

Despite improvements in the overall level of health and NCD burden in many European countries, significant regional inequalities remain. Our findings highlight these from an economic perspective, where the impact of productivity costs on regional GDP from cancer and CVD was relatively high in C-E Europe when compared to other European regions. The nature of this burden mirrors previous findings on geographical health inequalities, and is underpinned by a range of factors including disparities in socioeconomic levels, in addition to cultural differences impacting on lifestyle behaviours between eastern and western European countries [26]. A higher prevalence of smoking, higher overweight and obesity rates, and greater levels of hypertension are found in C-E Europe, which are all considered key risk factors for NCD [27].

The disparity in burden between cancer and CVD is also highlighted in our results, with a distinctly higher relative CVD productivity cost burden in C-E Europe compared to cancer. In addition to the common NCD risk factors noted above, the CVD burden also depends on the degree of disease control of hypertension, lipid, and glucose, for example [28]. Therefore, the higher relative CVD burden in C-E Europe is also, in part, a consequence of poorer access to early detection of CVD in addition to less investment in interventions for metabolic risk factors (lipid, glucose) and hypertension [28]. While previous studies have shown the potential health gains from reducing avoidable premature death especially in new European member states (predominantly CE European countries) [29], our study reveals the potential economic benefits of this for European economies, and for Central-Eastern European countries in particular, supporting arguments for further investment in interventions that have been shown to be previously cost-effective including early detection (for example, developing a register of patients who receive regular prophylactic penicillin) and treatment (for example, pharmacological and treatment of hypertension in adults) [30] in addition to a reduction in risk factors including tobacco and alcohol use, salt intake, raised blood pressure, lipid and glucose [31].

### Human capital and competitiveness aspects of the productivity cost burden in Europe

The loss to potential GDP from cancer and CVD is evidently sizable and is occurring against a backdrop of a stagnating European economy where waning competitiveness in the bloc has been highlighted, along with the importance of the quality and quantity of the labour force [32]. NCD deaths in the working age population reduces GDP and diminishes long term growth prospects through a detrimental impact on labour supply, productivity and capital investments [2–3]. This issue will become even more acute in the coming years given that the European population is experiencing rapid aging with more than a fifth now aged 65 years or older [33]. In 2022, there were 2·7 workers per elderly person, but moving forward, this is expected to fall to 1·5 by 2100 [33]. This demographic shift will increase the age-related burden of diseases such as CVD and cancer, imposing not only increasing pressure on limited public healthcare budgets, but also on economic growth and the quality of human capital, a development which can already be seen arising in this study where the estimated productivity costs were equivalent to 1·24% of the European GDP in 2021.

Regional inequality in productivity cost burden also emerged in our results, with C-E European countries experiencing a comparatively larger burden as a percentage of GDP. These countries are amongst the lowest per capita GDP countries in the region, and are expected to grow faster than the other regions (2.5-4%%) in the medium term [34]. This is necessary to raise living standards and lead to convergence in the European economy over time, however the human capital cost revealed in this study will continue to hinder the convergence process impacting the quality and quantity of the labour market in these countries. Western Europe also experienced a comparatively high burden as a percentage of GDP, highlighting again the aging population dynamics in these countries and the acute need to strategically invest in targeted NCD control initiatives for both health, and also future economic growth, purposes.

### Gender aspects of the productivity cost burden in Europe

A key finding from our results was that while both cancer and CVD exhibited higher male productivity costs compared to female (both total, and per death), CVD male productivity costs were proportionately greater (2·6 times versus 1·7). This disparity could be explained by the input of various epidemiological and economic factors in our analysis. Deaths from cancer and CVD tend to be higher among males than females, particularly CVD. This is exacerbated by higher wage levels and labour force participation rates for males which further adds to the gender disparity in the results. Interestingly, when we examined productivity cost per death, which adjusts for the difference in male and female mortality numbers, the male/female cost ratio decreases. This reflects the influence of economic factors on the gender disparity in the productivity cost burden and should be presented in future cost studies with a gender dimension. In addition, the application of gender-neutral wages reduced male/female cost ratios by between 14% and 16% across Europe for CVD and cancer, suggesting the importance of using gender-neutral wages in such costing exercises moving forward.

### Comparison with previous literature

Our productivity cost estimates for cancer and CVD are in excess of previous European estimates. For example, recent estimates for cancer across Europe were close to €50 billion in 2018 [10, 12] in comparison to our estimate of €122 billion. Similarly for CVD, previous estimates are less (€32·0 billion [9] versus 73·5 billion in our study). While some of the difference can be explained by the increasing number of deaths, there are two additional methodological reasons. Firstly, these previous studies capped costs at an assumed retirement age of 64, however, aging demographics are putting pressure on workers to continue to work beyond the traditional age of retirement across Europe and raising the cut off age to 74, where data shows that significant numbers of the European population are still working [15], as undertaken in this study, is a more realistic modelling stance. Indeed, cancer costs estimated in the US have traditionally taken a lifetime perspective or an older cut off age of 84, with premature mortality costs exceeding those estimated in Europe per death as a consequence. Secondly, further investigation revealed that traditional estimates do not appear to account for the potential lifetime wage progression of people who died from cancer or CVD. This paper adjusts wages in line with age and potential career progression had they not died prematurely. We would argue that this modelling approach more accurately reflects the true potential cost of NCDs to the economy.

### Strengths and limitations

Prior studies have independently assessed aspects of the CVD and cancer economic burden, focusing on direct healthcare costs, indirect costs (sometimes combined), and household financial consequences, but none do so for cancer and CVD productivity costs combined across Europe. To the best of our knowledge, this study is the first to directly compare the productivity costs associated with working age people with cancer and CVD in Europe using a comparable and standardised economic methodology. Our paper furthermore produces a more realistic estimate of productivity costs arising from cancer and CVD than previous reported studies by capturing wage-related age transition over the potential working lifecycle offering a new addition to the literature.

Our study does contain limitations, however. We did not attempt to capture the burden of unpaid work loss or direct healthcare costs as these were beyond the scope of the current study’s objective. We did not attempt to apply the friction cost approach as an alternative method of estimating productivity costs due to the onerous country-specific data requirements necessary for its successful application. We recognise the data limitations of using labour market data for individuals beyond 65 years of age where information is less reliable and assumptions were made by extrapolating data, and of attempting to apply past trends in the growth of economic variables to future estimates.

## Conclusions

Our results revealed the considerable magnitude of the combined cancer and CVD productivity cost burden amongst the working age population in Europe, and also highlighted regional differences in this burden with larger cancer costs across Southern, Western and Northern Europe, while CVD costs dominated in Central Eastern Europe. We expand the NCD burden map in Europe showing that cancer is responsible for a higher cost burden than CVD amongst populations of working age, supplementing the traditional health burden perspective. Our results further reveal the vast potential economic savings that could be generated, in addition to life expectancy gains, by reducing major behavioural risk factors for cancer and CVD.

## Data Availability

All data for this study are publicly available and can be found in World Health Organization. WHO Mortality Database. https://platform.who.int/mortality/themes/theme-details/MDB/all-causes and Eurostat. Database. https://ec.europa.eu/eurostat/data/database. These links are reproduced in the paper.

https://ec.europa.eu/eurostat/data/database

## Acknowledgements

None

## Supporting information Captions

**S1 Table: List of included countries by region and year of mortality data**

**S2 Table: Number of deaths for cancer and cardiovascular disease (CVD) among those aged 15-74 in Europe in 2021, overall and by sex.**

**S3 Table: Years of Potential Productive Life Lost for cancer and cardiovascular disease (CVD) among those aged 15-74 in Europe in 2021, overall and by sex.**

**S4 Table: Sensitivity analysis of total productivity costs for cancer and cardiovascular disease (CVD) by region, 2021.**

## Notes

### Competing Interest Statement

The authors have declared no competing interest.

### Funding Statement

The author(s) received no specific funding for this work.

